## SupplementaryFigure2 for "Comparative benchmarking of optical genome mapping and chromosomal microarray reveals high technological concordance in CNV identification and structural variant refinement"

### Slide 1
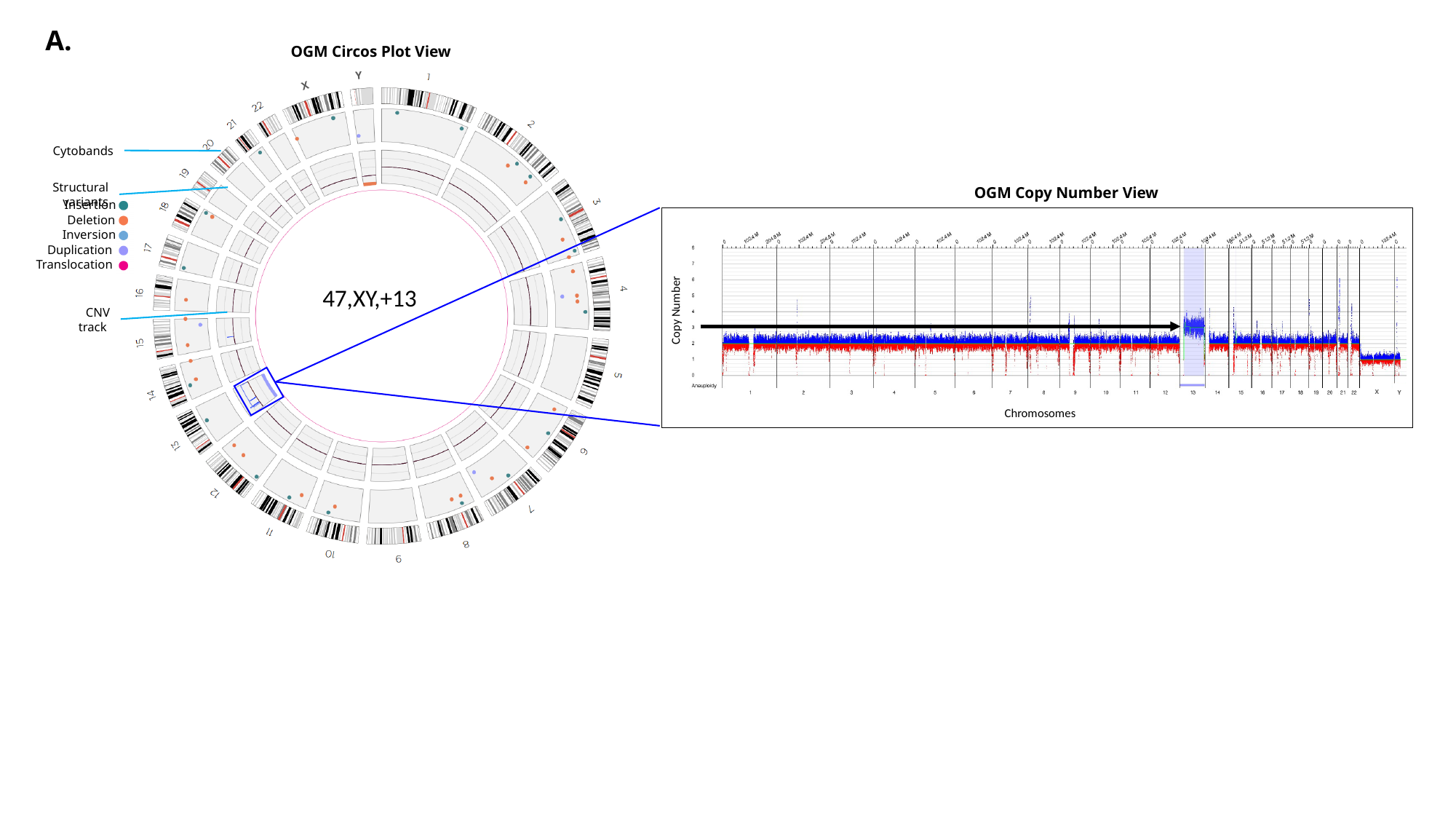

A.
OGM Circos Plot View
Y
X
Cytobands
Structural variants
OGM Copy Number View
Insertion
Deletion
Inversion
Duplication
Translocation
47,XY,+13
Copy Number
CNV track
X
Y
Chromosomes

### Slide 2
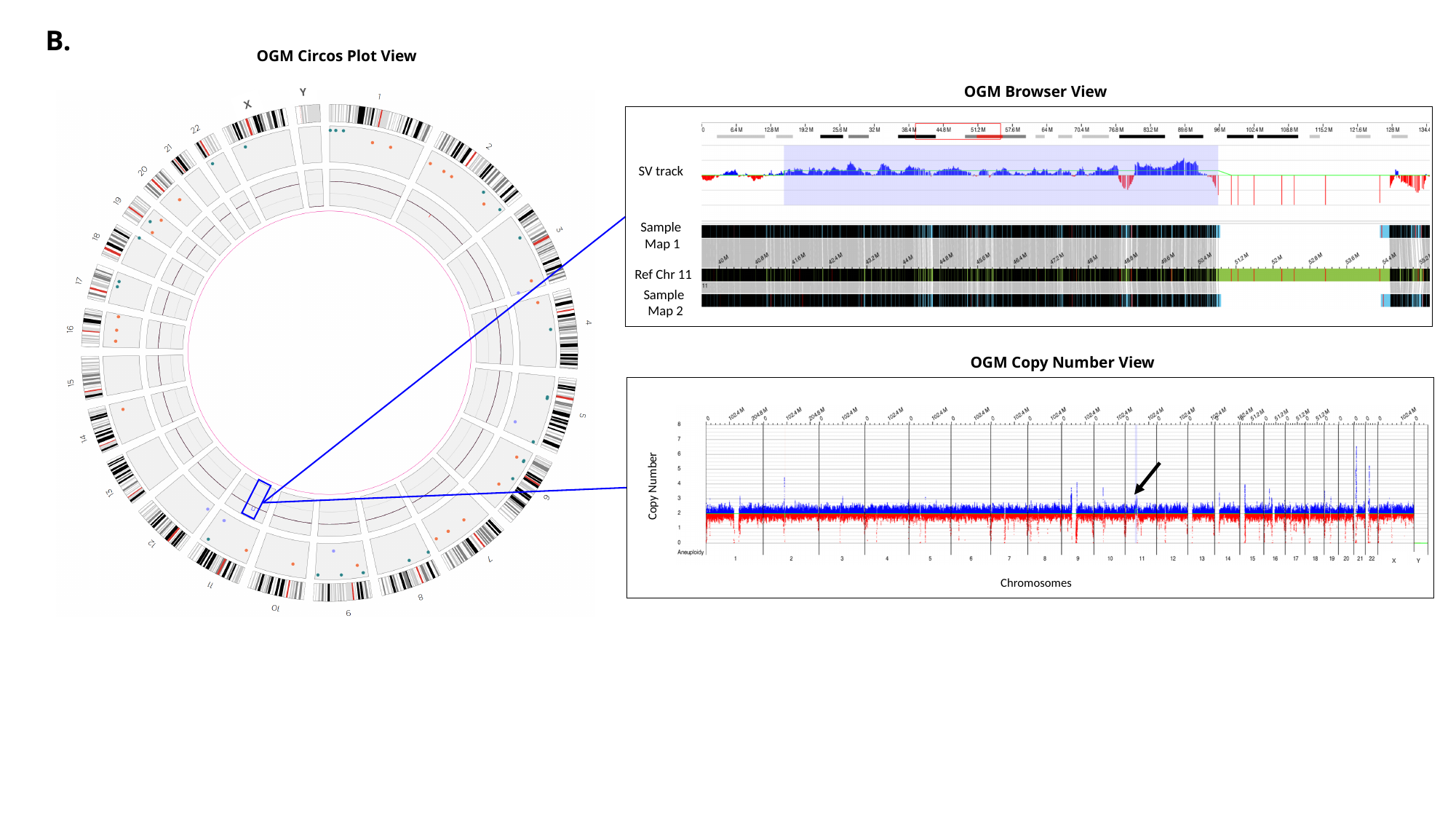

B.
OGM Circos Plot View
OGM Browser View
Y
X
SV track
Sample
Map 1
Ref Chr 11
Sample
Map 2
OGM Copy Number View
Copy Number
X
Y
Chromosomes

### Slide 3
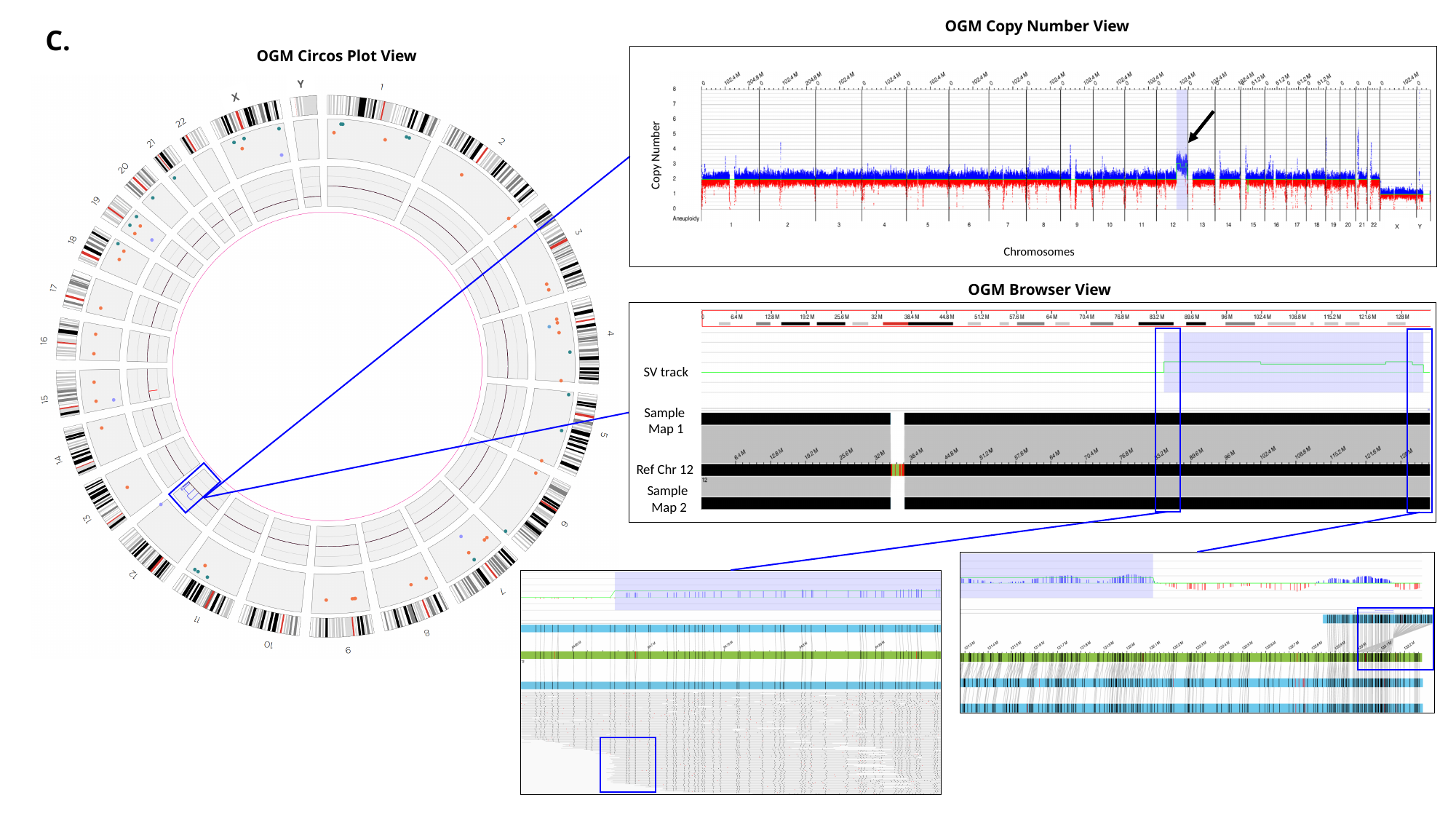

OGM Copy Number View
C.
OGM Circos Plot View
Y
X
Copy Number
X
Y
Chromosomes
OGM Browser View
SV track
Sample
Map 1
Ref Chr 12
Sample
Map 2

### Slide 4
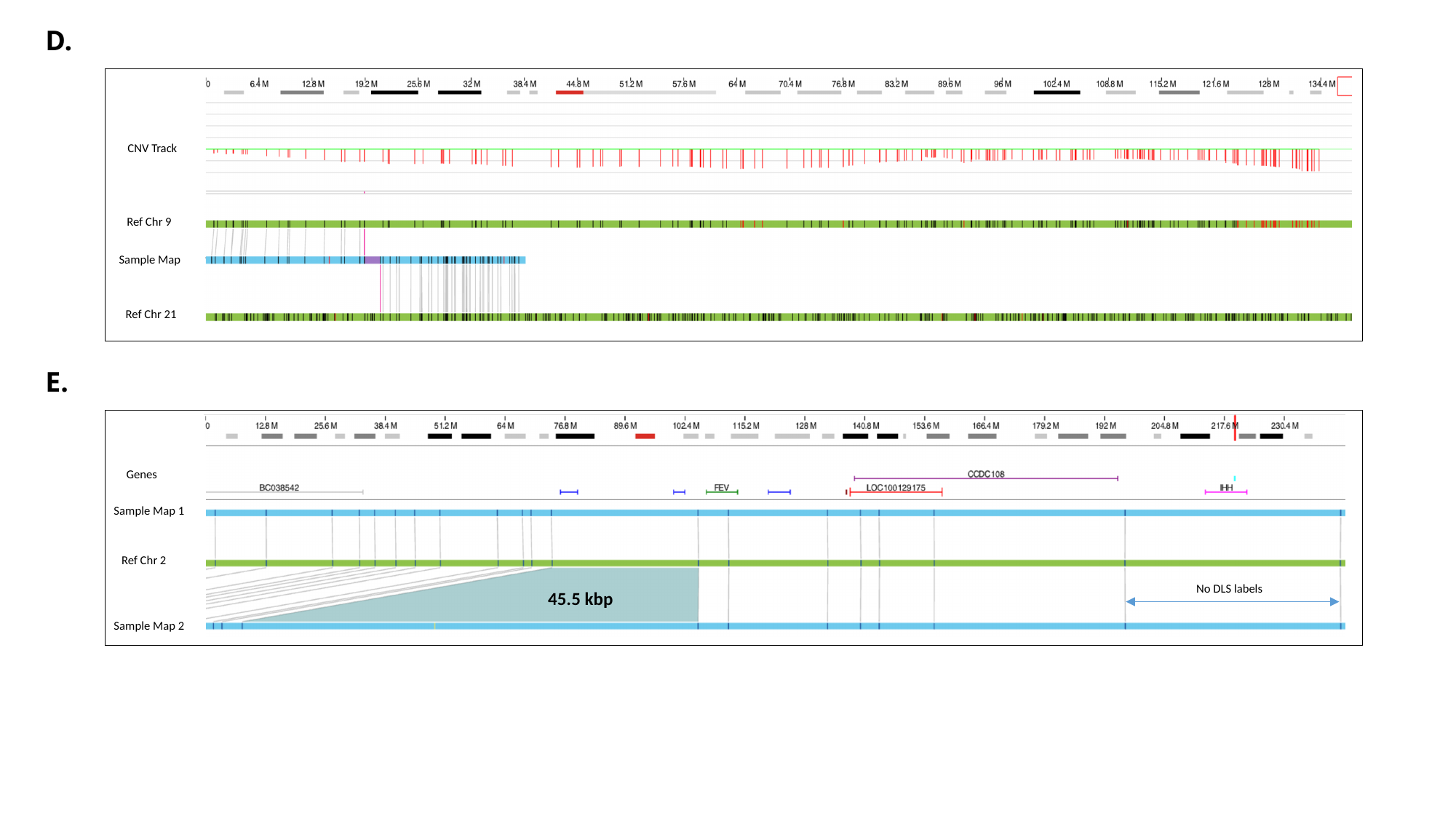

D.
CNV Track
Ref Chr 9
Sample Map
Ref Chr 21
E.
Genes
Sample Map 1
Ref Chr 2
No DLS labels
45.5 kbp
Sample Map 2
