## SupplementaryFigure1 for "Comparative benchmarking of optical genome mapping and chromosomal microarray reveals high technological concordance in CNV identification and structural variant refinement"

### Slide 1
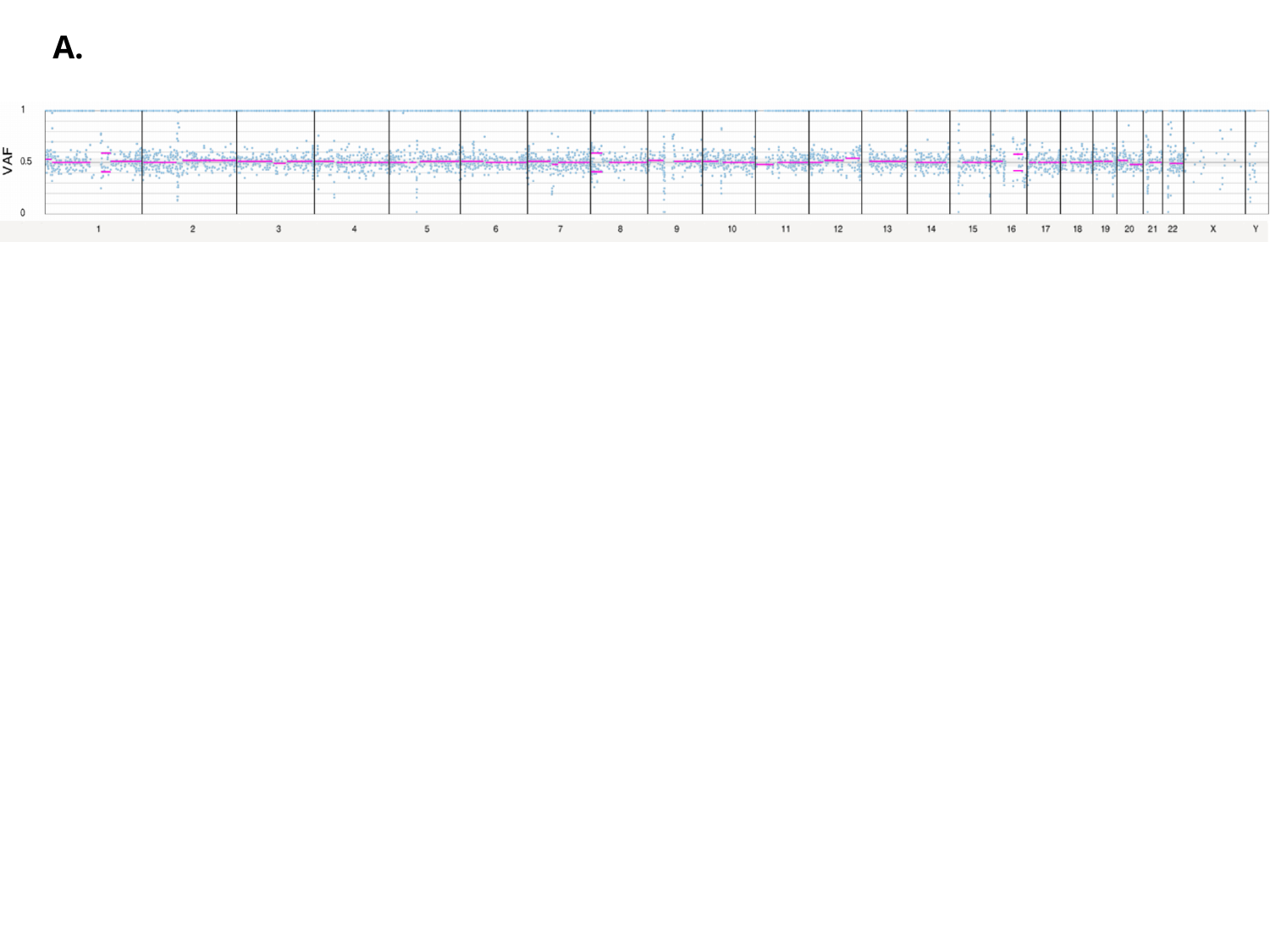

A.

### Slide 2
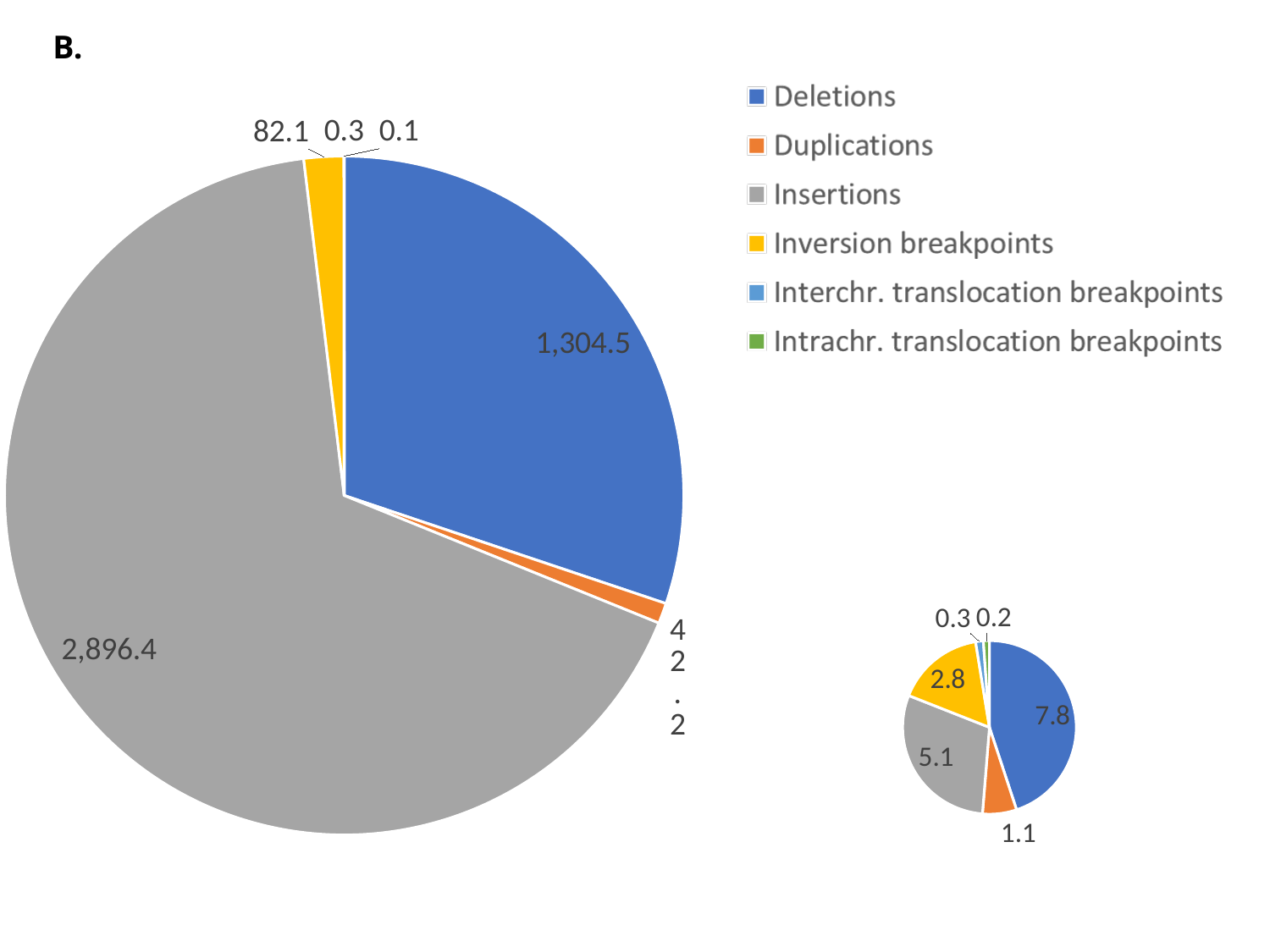

B.
#### Chart
| Category | |
|---|---|
| Deletions | 1304.5 |
| Duplications | 42.17307692307692 |
| Insertions | 2896.4423076923076 |
| Inversion breakpoints | 82.11538461538461 |
| Interchr. translocation breakpoints | 0.25 |
| Intrachr. translocation breakpoints | 0.1346153846153846 |
#### Chart
| Category | |
|---|---|
| Filtered deletions | 7.769230769230769 |
| Filtered Duplications | 1.0961538461538463 |
| Filtered Insertions | 5.134615384615385 |
| Filtered Inversion breakpoints | 2.8461538461538463 |
| Filtered Interchr. translocation breakpoints | 0.25 |
| Filtered Intrachr. translocation breakpoints | 0.19230769230769232 |

### Slide 3
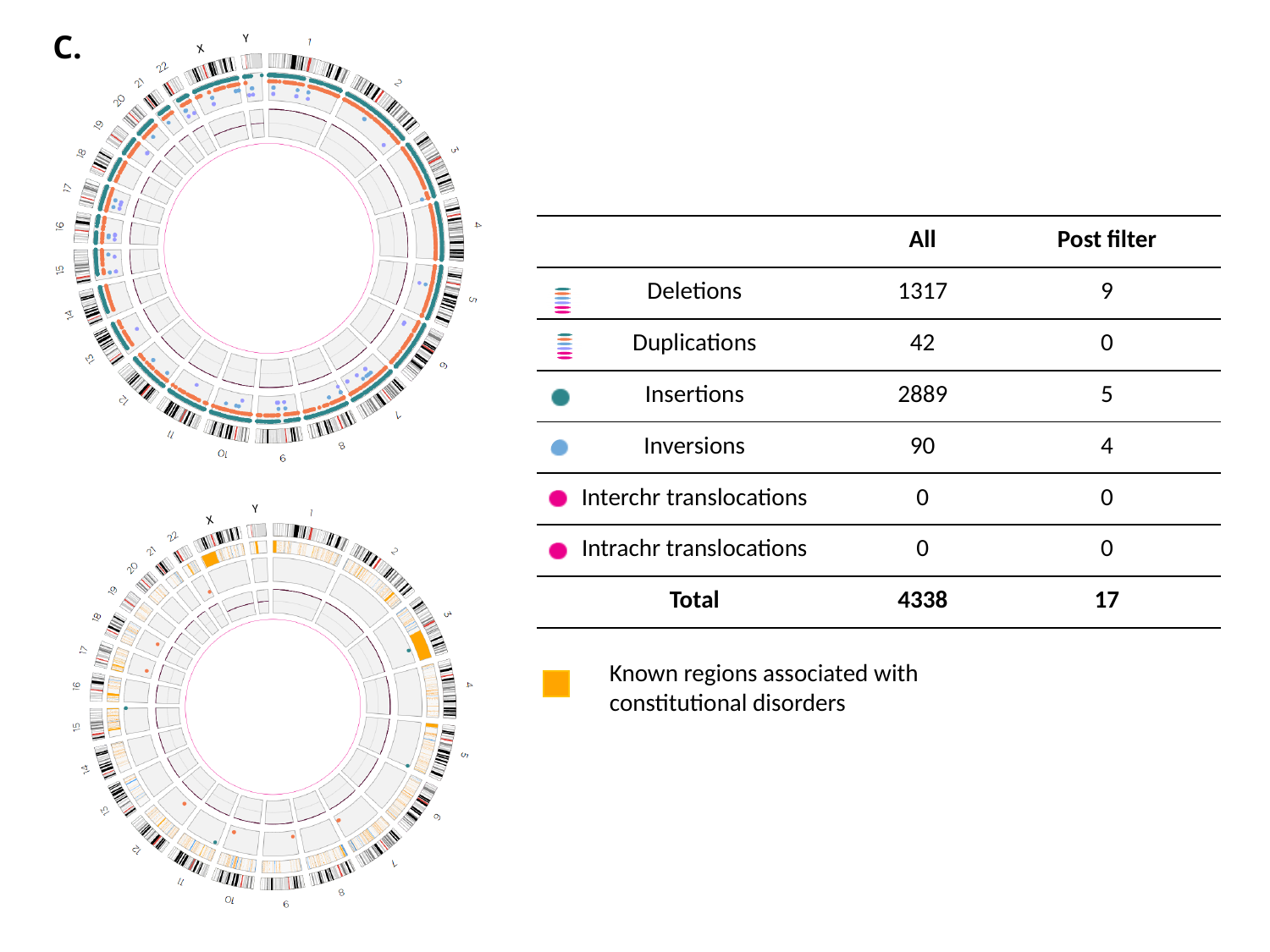

C.
Y
X
| | All | Post filter |
| --- | --- | --- |
| Deletions | 1317 | 9 |
| Duplications | 42 | 0 |
| Insertions | 2889 | 5 |
| Inversions | 90 | 4 |
| Interchr translocations | 0 | 0 |
| Intrachr translocations | 0 | 0 |
| Total | 4338 | 17 |
Y
X
Known regions associated with constitutional disorders
